# Associations of Alzheimer’s disease with inpatient hospital costs and with quality-adjusted life years: Evidence from conventional and Mendelian randomization analyses in the UK Biobank

**DOI:** 10.1101/2023.12.09.23299763

**Authors:** Padraig Dixon, Emma Anderson

## Abstract

**BACKGROUND:** Alzheimer’s disease and other dementias are progressive neurodegenerative disorders with profound impacts on cognitive function. There is a shortage of economic evidence relating to the impact Alzheimer’s disease on healthcare costs and quality-adjusted life-years (QALYs).

**METHODS:** We employed two study designs to model the association between Alzheimer’s disease and healthcare costs and QALYs. We first estimated conventional multivariable models of the association between Alzheimer’s disease and these core economic outcomes. However, these types of model may be confounded by diseases, processes, or traits that independently affect Alzheimer’s disease and either or both of healthcare costs and QALYs. We therefore also explored a complementary approach using germline genetic variation as instrumental variables in a Mendelian randomization analysis. We used single nucleotide polymorphisms (SNPs) identified in recent genome-wide association studies of Alzheimer’s disease as instruments. We studied outcome data on inpatient hospital costs and QALYs in the UK Biobank cohort.

**RESULTS:** Data from up to 310,838 individuals were analyzed. N=55 cases of Alzheimer’s disease were reported at or before recruitment into UK Biobank. A further N=284 incident cases were identified over follow-up. Multivariable observational analysis of the prevalent cases suggested significant impacts on costs (£1,140 in cases, 95% Confidence Interval (CI): £825 to £1,456) and QALYs (−25%, 95% CI: -28% to -21%). Mendelian randomization estimates were very imprecise for costs (£3,082, 95% CI: -£7,183 to £13,348) and QALYs (−32%, 95% CI: -149% to 85%), likely due to the small proportion of variance (0.9%) explained in Alzheimer’s disease status by the most predictive set of SNPs.

**IMPLICATIONS:** Conventional multivariable models suggested important impacts of Alzheimer’s disease on inpatient hospital costs and QALYs, although this finding was based on very few cases which may have included instances of early-onset dementia. Mendelian randomization was very imprecise. Larger GWAS of clinical cases, improved understanding of the architecture of the disease, and the follow-up of cohorts until old age and death will help overcome these challenges.

## 1 Introduction

Alzheimer’s disease, the most common cause of dementia, is a progressive neurodegenerative disorder that primarily affects cognitive function (1). Worldwide, some 57 million people were estimated to live with Alzheimer’s disease and other dementias in 2019 (2), a number that has more than doubled since 1990 because of population growth and population ageing (2)(3). The consequences of Alzheimer’s disease can be profound, especially as the disease progresses, and may include language impairment, personality changes, memory loss, and inability to manage daily tasks including self-care and rudimentary decision-making (1).

There are no cures for Alzheimer’s disease. Treatment of symptoms may stabilize cognitive function in some cases (4), but non-pharmacotherapeutic care and management remains essential for most patients. A key consideration for any intervention is cost-effectiveness, which is a function of the costs, quality-of life, and mortality impacts associated with both the disease and intervention under consideration. There have been relatively few economic evaluations of interventions for Alzheimer’s disease in comparison to other diseases, meaning that informed decisions are challenged by lack of evidence on how interventions affect these economics outcomes (5, 6). This type of evidence is particularly important for decision-analytic models of interventions for Alzheimer’s disease.

In this paper, we addressed this evidence gap by estimating associations of Alzheimer’s disease with healthcare costs and with quality-adjusted life years (QALYs) in the UK Biobank (7) cohort using two types of study design. We first explored the associations of Alzheimer’s disease with costs and QALYs in conventional multivariable models, which we henceforth simply refer to as multivariable models, using linear adjusted linear regressions that adjust for basic demographic information. Multivariable models are widely used as means of assessing associations between disease status and cost and QALY outcomes but may be confounded by omitted variables that influence both disease status and these outcomes. These confounding variables may include any processes or traits that independently influence Alzheimer’s disease and either or both of healthcare costs and QALYs.

We therefore also used germline genetic variation associated with the risk of Alzheimer’s disease as instrumental variables in a Mendelian Randomization. Mendelian Randomization exploits the natural experiment of quasi-random genetic variation allocation from parents to children during conception. Some of this allocation includes genetic variants known to influence the susceptibility to Alzheimer’s disease. This approach therefore avoids, in principle, confounding influences that might otherwise bias conventional multivariable models.

## 2 Data

Our principal data source was the prospective UK Biobank cohort (research ethics approval reference 11/NW/0382), which recruited over 500,000 people between 2006 and 2010 from 22 centres across England, Scotland, and Wales (6)(7–9).

### 2.1 Construction of cost and QALY data

We have previously described our creation of healthcare cost and quality-adjusted life years data (8-10). Briefly, the UK Biobank data was linked to medical records containing information on inpatient hospital episodes of care associated with diagnoses and procedures. From these data, we created per-patient, per year costs of inpatient hospital care from the linked Hospital Episode Statistics resource. We calculated costs from recruitment into Biobank up to the date of participant death or the censoring date for our Hospital Episode Statistics data of 31 March 2015.

QALY data were constructed by assigning quality of life decrements associated with 240 different health conditions (11) to the Biobank cohort. These specific health conditions were reported by individuals or recorded within their health records. This, in conjunction with data on mortality, facilitated the development of individual quality-adjusted life years (QALYs) from the time of recruitment until March 31, 2017. The individual, person-specific QALY estimates represent absolute percentage changes in QALYs over an average year of follow-up.

### 2.2 Identification of cases

Alzheimer’s disease cases in UK Biobank were defined as follows. Cases were identified as occurring at or before the baseline UK Biobank recruitment appointment if they were self-reported at that appointment, or if there was a prior record of inpatient care before the baseline appointment coded with ICD9 codes beginning with 331 or with ICD10 codes beginning with F00 (“Dementia in Alzheimer disease”) or G30 (“Alzheimer disease”). The self-report question was not specific to Alzheimer’s disease but instead queried history of “Dementia/Alzheimers/Cognitive Impairment” Only cases identified at or before the baseline recruitment appointment were used in the analysis to ensure that the exposure was known to have happened before outcomes were observed. In addition to these cases, we also identified incident cases from either hospital records (using the ICD codings above) or from self-report at a post-baseline interview to which some participants were invited. (23). For the Mendelian randomization analysis, we considered both prevalent (known at or before baseline) and incident cases (occurring after baseline recruitment) since the exposure variable was PRSs instrumenting lifelong genetic liability to Alzheimer’s disease. The multivariable analysis considered only Alzheimer’s disease reported at or before baseline to ensure the cost and QALY outcomes were observed after exposure was recorded.

### 2.3 Polygenic risk scores for Alzheimer’s disease

We created weighted polygenic risk scores (PRSs) from genome-wide association studies (GWASs) for the two-sample Mendelian randomization analysis. We used Alzheimer’s disease GWASs that did not use data from the UK Biobank cohort (our outcome data source) in order to avoid bias from sample overlap (12). We examined two recent GWASs in individuals of European ancestry with clinically confirmed Alzheimer’s disease or dementia: Kunkle et al (13) and Bellenguez et al (14). The Kunkle et al GWAS was published in 2019 and analysed data from 63,296 individuals in a discovery sample with clinically diagnosed late-onset Alzheimer’s disease. Bellenguez et al was published in 2022 and used a two-stage design with up 487,511 individuals included in discovery sample analysis comprising both diagnosed and proxy cases (based on whether respondents reported their parents having dementia), although we only used results from the GWAS of clinically diagnosed cases in our analyses.

We used an in-house protocol (15) to quality-control genetic data from UK Biobank. We applied an R^2^ threshold of 0.001 within a 10,000-kilobase window to clump genome-wide significant SNPs to minimize linkage disequilibrium. For SNPs not present in the UK Biobank outcome dataset, we conducted a search for proxy SNPs using the European subsample of the 1,000 Genomes Project as a reference panel, setting a lower R^2^ limit of 0.6 (11). We restricted the main analysis to unrelated individuals of white British ancestry (to avoid confounding by ancestral background) living in England or Wales at recruitment, and after removal for sex mismatches, sex chromosomal abnormalities, and withdrawals of consent.

Using the set of SNPs remaining after this process, PRSs were calculated as the weighted sum of the risk-increasing effect alleles of genome-wide significant SNPs, with each SNP’s weight determined by the log-odds ratio coefficient obtained from the corresponding GWAS. We proxied variation explained in Alzheimer’s disease by the PRS using Nagelkerke’s pseudo R^2^ calculated from a logistic regression of Alzheimer’s disease case status on each respective PRS, as well as the R^2^ adjusted for the effective sample size in these logistic regressions, which is a function of the proportion of observations at each distinct value of the Alzheimer’s disease case status variable.

We also examined the impact of including two variants from the APOE (Apolipoprotein E) region, encoded by the rs7412 and rs429358 SNPs. The APOE ε2 allele (encoded by the rs7412 SNP) is protective for incident Alzheimer’s disease, the APOE ε4 allele (encoded by rs429358) increases risk, and together the interplay between these two SNPs exerts an important influence on genetic susceptibility to Alzheimer’s disease (16, 17). We therefore examined the impact of excluding this pair of SNPs from each analysis and assessed how well these two SNPs alone explained disease-case status.

### 2.4 Data and code availability

The derived cost and QALY variables will be archived with UK Biobank as returned variables and will be made available to individuals who obtain the necessary permissions from the study’s data access committee. Analysis code is available here: https://github.com/pdixon-econ/alzheimer-cost-qaly.

## 3 Methods

We estimated conventional multivariable linear regressions for QALYs and healthcare costs in relation to Alzheimer’s disease status, with age, sex, 40 genetic principal components, Townsend Deprivation Index, and recruitment centre as controls. We also re-ran these regressions separately for men and women. All multivariable analyses were restricted to the same group of individuals for which we could perform the Mendelian Randomization analysis, which comprised individuals who gave consent to be genotyped, and who passed the quality control and processing steps described below. The multivariable analysis differed from the Mendelian Randomization analyses by studying only prevalent cases (reported before or at recruitment into UK Biobank) to ensure exposure to Alzheimer’s disease occurred before the outcomes were observed.

We then undertook the Mendelian Randomization analysis (18-21). Under three core assumptions, Mendelian Randomization may be used to estimate the causal effect of an exposure on an outcome. The first of these assumptions is that the instrument be associated with the exposure of interest. The genetic variants used as instruments in MR are typically single nucleotide polymorphisms (SNPs), which represent single nucleotide changes in the genetic code. SNPs are a type of allele; alleles are the alternative forms of a gene at a particular location in the genome. The associations of specific SNPs with disease risk are typically tested using genome-wide association studies. The second assumption, sometimes referred to as the independence requirement, is that there is no confounding of the instrument/outcome relationship. The third assumption, often known as the exclusion restriction, is that the instrument affects the outcome only via the exposure. These may include other diseases, traits or behaviours, or may involve processes such population structure (since ancestral background influences both the distribution of genetic variants and also disease risk not necessarily associated with those variants) and geographic structure (since individuals with the same or similar ancestral background living in different types of location may experience different environmental influences that affect the risk of Alzheimer’s disease).

Two other important influences may also violate these assumptions. First, horizontal pleiotropy – the influence of a single variant on multiple phenotypes – may violate the exclusion restriction assumption if a variant influences the outcome by paths that are independent of the exposure. Second, linkage disequilibrium, which refers to the non-random association of alleles at different genomic loci to be inherited together. This could lead to the association of a variant included in analysis with excluded variants, potentially violating the third assumption.

Neither the second nor the third assumption is fully testable, but potential violations can be identified. Below, we describe the modelling approaches and data processing steps we took to account for possible failures of the core Mendelian Randomization assumptions. If the assumptions are satisfied, the analysis will identify groups that have varying levels of exposure to the risk of Alzheimer’s disease and for which outcomes – costs and QALYs – may be compared.

### 3.1 Polygenic risk scores

For the Mendelian randomization models, we implemented two-stage least square (2SLS) instrumental variable models. We first regressed the Alzheimer’s disease case status variable on the PRS, adjusting for age at baseline recruitment in UK Biobank, the first forty genetic principal components (as a control for population structure), and the UK Biobank recruitment location (as a control for geographic structure). Predicted values from this first stage regression were then used in a second stage regression on the cost and QALY outcomes respectively. 2SLS models were estimated using the AER package (22) in R. We inspected F-statistics from the first-stage regression to evaluate instrument strength (23).

The effect estimates from these PRS models reflect genetic liability to disease, and may be interpreted as the average effect (on either costs or QALYs) per unit increase in the log-odds of having Alzheimer’s disease (23, 24). This assumes either a constant effect of genetic liability to Alzheimer’s disease on the outcomes, or a monotonic effect of the SNPs on Alzheimer’s disease risk under a local average treatment effect interpretation.

### 3.2 Pleiotropy-robust sensitivity analysis

We also examined potential violations of the exclusion restriction assumption due to horizontal pleiotropy. We examined heterogeneity in effect estimates across individual SNPs using Cochran’s Q statistic (25). Heterogeneity may be due to sampling error, or to the presence of SNPs with a large influence (relative to the mean effect of all SNPs) on outcome, which could suggest a channel or channels of influence that go from the SNP to the outcome other than via Alzheimer’s disease. This latter form of heterogeneity would indicate a likely violation of the exclusion restriction.

In our main analysis, we calculated IVW estimates for individual SNP instrumental variables, combining them via random-effects meta-analysis with weights based on SNP-outcome precision. This assumes no exclusion restriction violations, or restrictions that affect only the variance of estimates but not the overall effect size. We then implemented three estimators robust to more general forms of pleiotropy, each with their own assumptions: the MR Egger estimator, which is consistent under assumptions related to instrument strength and pleiotropic effects (14, 15); a penalized weighted median estimator, consistent if ≥50% SNPs are valid instruments (16); and a weighted mode estimator, consistent if the largest homogenous SNP cluster is valid, even if more than 50% SNPs are invalid instruments (17).

## 4 Results

We analysed data from data from up to 310,838 unrelated individuals in multivariable and Mendelian randomization analyses, of whom 53.5% were female. Mean age at recruitment in this sample was 56.9 years. Cost data were available for a median of 6.1 years of follow-up and QALY data for a mean of 8.1 years. Mean healthcare costs per person, per year of follow-up were £88 in 2019 prices and mean per-person QALYs (averaged over all years of follow-up) were 0.91.

There were 55 cases of Alzheimer’s disease self-reported at or before baseline, and 339 cases including these cases as well as incident diagnoses recorded in inpatient care records over the follow-up. Thus just 0.1% of the sample had Alzheimer’s disease before or during UK Biobank recruitment and follow-up, although 12.6% (n=39,195) of the sample reported that at least one of their parents had Alzheimer’s disease.

A PRS comprised of 22 SNPs based on Kunkle et al (13) (including the rs7412 and rs429358 SNPs in the APOE locus) was the most predictive of the PRSs tested, with a Nagelkerke pseudo-R^2^ of 0.9% and an R^2^ adjusted for effective sample size of 3.9%. Results for other PRSs tested are given in Supplementary material (Table S1) and range from <0.0% to 0.9% for Nagelkerke’s R^2^ and <0.0% to 3.9% for R^2^ adjusted for effective sample size. This 22-SNP PRS was associated with prevalent and incident Alzheimer’s disease (log odds-ratio 0.57, 95% CI: 0.41 to 0.74) and with self-reported parental history of Alzheimer’s disease (log odds-ratio 0.25, 95% CI: 0.23 to 0.27), but there was no evidence of an association with the self-reported baseline cases (log odds-ratio 0.16, 95% CI:-0.30 to 0.62). This latter finding could be a consequence of the non-specific self-reported question asked at baseline in UK Biobank, instances of early-onset dementia assessed against a PRS for late-onset disease, or low statistical power given the small number of cases.

### 4.1 Results of main analysis

The multivariable analysis indicated material increases in cost and decreases QALYs associated with self-reported cases of Alzheimer’s disease at baseline. These associations were similar for men and women (Table 1) with overlapping confidence intervals, although point estimates for men indicated more severe outcomes for both costs and QALYs.

**Table 1.** Adjusted multivariable estimates of association between self-reported Alzheimer’s disease and costs and QALYs.

|  | N | Effect estimate | 95% confidence interval |
| --- | --- | --- | --- |
| **Healthcare costs** |  |  |  |
| All | 310,496 | £1,140 | £825 to £1,456 |
| Men | 143,841 | £1,281 | £851 to £1,710 |
| Women | 166,655 | £906 | £432 to £1,380 |
| **QALYs** |  |  |  |
| All | 310,496 | -25% | -28% to -21% |
| Men | 143,841 | -27% | -32% to -22% |
| Women | 166,655 | -21% | -26% to -15% |

These models adjust for age, sex, the Townsend deprivation index, and UK Biobank recruitment centre. Note that Townsend deprivation index was not available at baseline for all Biobank participants and the N of these regressions is therefore smaller than the total sample size.

The Mendelian randomization estimates were very imprecise, being consistent with both large and small impacts of Alzheimer’s disease on both costs and QALYs (Table 2). The uncertainty around point estimates means that conclusions about the potential magnitudes of these impacts is not possible. Given this uncertainty, we did not disaggregate results by sex.

**Table 2.** 2SLS PRS estimates.

|  | **Effect estimate** | **95% confidence interval** |
| --- | --- | --- |
| **Outcome** |  |  |
| Healthcare costs | £3,082 | -£7,183 to £13,348 |
| QALYs | -32% | -149% to 85% |

The F-statistic was 47.8 for both costs and QALY outcomes (since the first stage regression in each case is identical). While this is above the conventional F>10 rule of thumb sometimes used in this context, weak instrument bias relates to instrument strength in a continuous rather than binary manner and we cannot rule out the possibility of bias. If present, weak instruments would bias these Mendelian randomization estimates in the direction of the conventional multivariable estimates.

### 4.2 Sensitivity analysis

Cochran’s Q test for heterogeneity was consistent with the null for the cost outcome (Q=13.3, p=0.82) but there was evidence of heterogeneity in the QALY estimates (Q=39.8, p<0.01). Inspection of the IVW and various pleiotropy robust estimators (MR Egger, penalized weighted median, and weighted mode) revealed similar point estimates for the QALY outcome but these were consistent with the null in all cases and associated with considerable uncertainty (See Supplementary Material Table S3).

## 5 Discussion

We examined the association between Alzheimer’s disease and the core health economic outcomes of healthcare cost and QALYs using two types of study design in the large UK Biobank cohort. Our analysis was limited to data from individuals of European ancestry. Conventional multivariable estimates indicated large impacts on each outcome when assessed using cases identified at or before recruitment into the UK Biobank cohort. However, the number of such cases was very small relative to the size of the cohort. Moreover, these cases may have included instances of early onset dementia, although we cannot establish if this the case.

We also estimated Mendelian randomization models using genome-wide significant genetic variants from Kunkle et al. While this type of analysis may be less biased by confounding and reverse causation than the multivariable models, the results of the Mendelian randomization analysis (both the 2SLS models and the pleiotropy robust models) were too imprecise to draw meaningful conclusions about the magnitude of Alzheimer’s disease impacts on these outcomes. This uncertainty was likely driven by the small number of cases in the UK Biobank cohort, and the modest explanatory power of the best performing polygenic risk score analysed.

This is the first Mendelian randomization analysis of the causal effect of Alzheimer’s disease on QALYs and any form of healthcare cost. The literature assessing and evaluating findings from more conventional study designs against these outcomes typically reports very large cost impacts e.g. (26-31). The 1999 work by Neumann et al (32) remains a heavily used source of evidence on quality-of-life associated with this condition, finding that disease status was a significant predictor of health-related quality-of-life scores amongst patients, with negative impacts increasing in the severity of the condition. A recent systematic review and meta-analysis (32) of health-related quality-of-life also found this dose-dependent relationship over disease severity. Point estimates were broadly comparable to the point effects obtained in our multivariable and Mendelian Randomization estimates.

Our cost data was limited to inpatient costs incurred in NHS hospitals linked to the Biobank cohort. We did not model outpatient or other healthcare costs; for example, we could not account for care provided in outpatient neurology clinics. Moreover, inpatient costs are likely to be much higher in the advanced stages of disease and towards the end of life; thus, we may not fully capture the extent of those in a relatively young cohort like the UK Biobank. Total healthcare costs associated Alzheimer’s disease are very likely to be higher than those we report here, although in principle the Mendelian randomization estimates may offer an unbiased estimate of exposure to the condition on inpatient hospital costs for this age range.

We also did not have access to carer costs, which are a very significant issue for most Alzheimer’s disease patients and their families (33). While research continues on the most appropriate way to account for carer burden in economic evaluation (34, 35), further evidence on the scale of potential impacts in this group is needed (36-39).

We generated QALYs by assigning quality of life decrements to 240 specific health conditions reported in episodes of inpatient hospital care, amongst which conditions were Alzheimer’s disease and other dementias. This created a modest degree of circularity, since case status therefore appeared in both the exposure and outcome. However, the overall contribution of case status to variability in the QALY outcome was very small given the low number of such cases recorded in UK Biobank. Our indirect calculation of quality-of-life data avoided complexities associated with declining ability to self-complete quality-of-life questionnaires as the disease progresses (40). The QALY calculation was based on conditions reported in episodes of hospital care and this may have overstated quality-of-life at baseline by considering only inpatient care.

Two types of selection bias may have affected both types of analysis. The first is that UK Biobank is a volunteer cohort, which relied on participants being sufficiently healthy and motivated to participate. While Alzheimer’s disease primarily manifests in later life, very unwell individuals, including with early or prodromal disease, would have been less likely to participate (41). UK Biobank participants are known to be healthier (on average) compared to the wider population from which the cohort was drawn (31). Even if this type of selection related to morbidity and survival was not present or not material (given the late onset of most cases of Alzheimer’s disease), the implicit conditioning on participation may also amount to survival or collider bias (42). This would arise if factors related to Alzheimer’s disease and the outcomes (such as high healthcare costs or low quality-of-life driven by significant morbidity) themselves influenced participation, which could confound the associations that we present in both multivariable and Mendelian randomization models.

The Mendelian randomization estimates reflect the impact of lifelong exposure to elevated genetic liability to incident Alzheimer’s disease, whereas the multivariable estimates reflect both this lifelong exposure as well as possible confounding influences from other variables. Neither the multivariable nor the Mendelian randomization estimate necessarily correspond to how outcomes would change under modification of Alzheimer’s disease risk, including via hypothetical manipulation of disease risk factors. Nevertheless, Mendelian randomization estimates likely reflect many of the common germline influences on genetic risk for this condition.

Our analysis has highlighted several areas for future work. There remains a need for further, larger GWASs of clinically diagnosed Alzheimer’s disease (43, 44). Improved understanding of the genetic basis of the disease will support aetiology more generally, as well as downstream causal analysis utilizing Mendelian randomization. The Mendelian randomization analysis focused on relatively common SNPs; future analysis could explore the impact of rare variants with large effects on disease risk (43). UK Biobank aimed to recruit individuals aged between 40 and 69, and linked care records enable longitudinal aspects of disease incidence and progression to be evaluated as the cohort ages. However, recruiting older individuals to future cohorts, as will continued study of cohorts such as the various international longitudinal studies of ageing, will comprise an important element of future research in this area.

## 6 Conclusion

Alzheimer’s disease is associated with significant impacts on the core health economic outcomes of (inpatient hospital) costs and QALYs. However, these associations were estimated with considerable uncertainty in causal analyses. Further interrogation and understanding of the genetic architecture of Alzheimer’s disease, and on the wider costs and quality-of-life impacts (including at the more advanced stages of disease) of the condition, will shed further light on these impacts.

## Data Availability

The derived cost and QALY variables will be archived with UK Biobank as returned variables and will be made available to individuals who obtain the necessary permissions from the study`s data access committee. Analysis code is available here: https://github.com/pdixon-econ/alzheimer-cost-qaly.

## Acknowledgments

This research was conducted using the UK Biobank Resource under Application Number 29294.

