## Supplementary material for "Associations of Alzheimer’s disease with inpatient hospital costs and with quality-adjusted life years: Evidence from conventional and Mendelian randomization analyses in the UK Biobank"

### 1 Performance of different polygenic risk scores

We explored the proportion of variance explained in our Alzheimer's disease phenotype (including prevalent and incident cases) using unadjusted logistic regression with disease status as the outcome and different polygenic risk scores (PRSs). We examined Nagelkerke's pseudo-$R^2$ statistic as well as $R^2$ adjusted for effective sample size. We used the score that explained most variance in the 2SLS regressions that are reported in the main text.

We extracted genome-wide significant SNPs from Kunkle et al (1) and for rs7412 and rs429358 from the IEU GWAS database. We extracted SNPs from Supplementary Table 7 of Bellenguez et al (2), and included only clinically confirmed cases in the sample which excluded UK Biobank. In all cases, the SNPs are those remaining after processing by in-house protocols (including searches for proxy SNPs in UK Biobank) described in Mitchell et al (3). The lower $R^2$ for the APOE SNPs when combined with the Bellenguez et al SNPs compared to the $R^2$ for these SNPs alone is possibly explained by interactions amongst the SNPs, although we did not attempt to fully account for this finding.

**Table S1 $R^2$ of different polygenic risk scores**

| | N of SNPs | Nagelkerke's $R^2$ | $R^2$ adjusted for effective sample size |
|---|---|---|---|
| **Polygenic risk scores** | | | |
| Kunkle et al incl rs7412 & rs429358 | 22 | 0.9% | 3.9% |
| rs7412 & rs429358 only | 2 | 0.2% | 1.1% |
| Bellenguez et al | 82 | <0.0% | <0.0% |
| Bellenguez et al w/ rs7412 & rs429358 | 84 | 0.1% | 0.3% |

### 2 SNPs included in the PRS model used in the 2SLS Mendelian Randomization analysis

**Table S2 Included SNPs**

| SNP | Chr | Beta (log OR) | Std Error | P-value | Effect allele | Other allele |
|---|---|---|---|---|---|---|
| rs1081105 | 19 | 0.942 | 0.0436 | 1.51E-103 | C | A |
| rs111278137 | 19 | 0.4735 | 0.0713 | 3.20E-11 | G | A |
| rs11257242 | 10 | 0.0841 | 0.0154 | 4.64E-08 | G | C |
| rs114812713 | 6 | 0.298 | 0.0431 | 4.47E-12 | C | G |
| rs11767557 | 7 | 0.1028 | 0.0182 | 1.56E-08 | T | C |
| rs12151021 | 19 | 0.1071 | 0.0169 | 2.56E-10 | A | G |
| rs12590654 | 14 | 0.0906 | 0.0157 | 8.73E-09 | G | A |
| rs139136389 | 19 | 0.4938 | 0.0851 | 6.43E-09 | C | T |
| rs147711004 | 19 | 1.1354 | 0.0366 | 1.00E-200 | A | G |

| | | | | | | |
|---|---|---|---|---|---|---|
| rs150685845 | 19 | 0.5561 | 0.0645 | 6.62E-18 | G | A |
| rs1582763 | 11 | 0.1232 | 0.0149 | 1.19E-16 | G | A |
| rs34665982 | 6 | 0.0967 | 0.0166 | 5.80E-09 | T | C |
| rs3740688 | 11 | 0.0935 | 0.0144 | 9.70E-11 | T | G |
| rs3851179 | 11 | 0.1198 | 0.0148 | 5.81E-16 | C | T |
| rs6733839 | 2 | 0.1693 | 0.0154 | 4.02E-28 | T | C |
| rs679515 | 1 | 0.1508 | 0.0183 | 1.55E-16 | T | C |
| rs72654445 | 19 | 0.5425 | 0.0811 | 2.27E-11 | G | A |
| rs73223431 | 8 | 0.0936 | 0.0153 | 8.34E-10 | T | C |
| rs7412 | 19 | 0.4673 | 0.0305 | 6.40E-53 | C | T |
| rs867230 | 8 | 0.1333 | 0.0158 | 3.49E-17 | A | C |
| rs9381563 | 6 | 0.0821 | 0.0148 | 2.93E-08 | C | T |
| rs429358 | 19 | 0.183737 | 0.0189 | 1.00E-200 | C | T |

### 3 Pleiotropy-robust sensitivity analysis

This section presents the results of the two-sample summary Mendelian Randomization sensitivity analyses for the cost and QALY outcomes. The effect estimates cannot be directly compared to the Mendelian Randomization 2SLS results presented in Table 2 in the main paper. The 2SLS results may be interpreted as the impact of genetically influenced change in case status, whereas the effect estimates presented below are on the logistic scale of the Kunkle et genome wide association study. The effect estimates in Table S4 therefore correspond to the change in costs or the change in QALYs per unit change in the log-odds of developing Alzheimer's disease.

**Table S3 Results of Inverse variance weighted and pleiotropy robust Mendelian Randomization estimators**

| | Estimate | Standard error | P-value |
|---|---|---|---|
| **Costs** | | | |
| Inverse variance weighted | £3 | £4 | 0.46 |
| MR Egger | £4 | £6 | 0.49 |
| Penalized weighted median | £0 | £5 | 0.98 |
| Weighted mode | -£1 | £6 | 0.89 |
| **QALYs** | | | |
| Inverse variance weighted | -0.03% | 0.06% | 0.69 |
| MR Egger | -0.1% | 0.1% | 0.31 |
| Penalized weighted median | -0.04% | 0.06% | 0.49 |
| Weighted mode | -0.03% | 0.07% | 0.71 |
